## Supplementary material for "A Novel Application of Neural Networks to Identify Potentially Effective Combinations of Biologic Factors for Enhancement of Bone Fusion/Repair": OrthoBioSupplement

This supplement contains items that support the analysis presented in the above-referenced article, the purpose of which is to identify combinations of factors that would provide better outcomes in bone repair than the factors currently in use. The core idea is to train an artificial intelligence (AI) using machine learning on available data, and then use the AI to predict the efficacy of many combinations of factors and identify the potentially most effective combinations. Clinical evaluation of the best combinations would justifiably follow their experimental verification.

**Text S1: Setting up the training data**

Available data were curated from the published literature and organized into a format suitable for machine learning. When possible, metanalyses providing tables listing outcomes for specific therapeutic trials were used for convenience of data collection. When metanalyses did not include appropriate data in table format, specific articles were used to obtain intervention (input) and outcomes (output) data. Literature used specifically to compile the training data can be found at the end of this supplement.[1-12]

After obtaining input and output data, the data were organized into input/desired-output pattern pairs. The inputs were the following medical interventions, with dosage/coding specifications in parentheses: low intensity pulsed ultrasound (total days of post-operative treatment), electrical stimulation (utilization post operatively (1) versus no utilization (0)), rhBMP2 (milligrams), rhBMP2 ND (no dose specified, utilized (1) versus not utilized (0)), rhBMP7 (milligrams), autogenous bone graft (utilization intra-operatively (1) versus no utilization (0)), exogenous bone graft (utilization intra-operatively (1) versus no utilization (0)), gelatin-calcium sulfate hydroxyapetite biomaterial (utilization intra-operatively (1) versus no utilization (0)), zoledronic acid (micrograms), osteogenin (micrograms), platelet-derived growth factor (micrograms), platelet-derived growth factor (coated, rat model), bone marrow aspirate concentrate (utilization intra-operatively (1) versus no utilization (0)), human (not human study (0) versus human study (1)), rat (not rat model (0) versus rat model (1)), animal model (not animal model other than rat (0) versus animal model other than rat (eg mouse, horse) (1)), platelet rich plasma (micrograms). Osteoconductive agents (and other bone scaffolds) were included in some studies described in the literature we accessed but we excluded them from the data set except for allograft, as the goal of our study was to characterize biologic factors which could be used in combination within an allograft framework.

The following outputs were outcome measures from clinical and basic science trials, with dosage/coding specifications in parentheses: distraction rate (millimeter per day), bone formation at 3 months (percentage of sample with positive bone formation), bone formation at 6 months (percentage of sample with positive bone formation), mineralized tissue volume divided by total tissue volume, 1-level posterior lumbar fusion percentage (fusion rate percent in sample), Oswestry disability index (ODI) improvement (percentage of sample with an improvement in ODI), fusion rate (percentage of sample with complete bone fusion), fracture healing rate (percentage of sample with complete fracture healing), time to achieve full weight bearing or clinical healing (in days), mean time to radiographic union (in days), Oswestry score (range 0-100), patients satisfied (percentage of patients who are “satisfied” with outcome), need for repeat bone grafting (number of patients), not healed at end of trial (number of patients), need for dynamization (number of patients), radiographic outcome score (assessed by radiologist/orthopedist), histomorphometric/histologic outcome score, implant survival (percent of sample with implant survival), radiopacity (percent of sample with radiopaque bone on radiograph), new bone formation (in $mm^2$), subsidence (complication occurred (1) versus no complication (0)), graft malposition/loosening (complication occurred (1) versus no complication (0)), infection (complication occurred (1) versus no complication (0)), urogenital adverse events (complication occurred (1) versus no complication (0)), retrograde ejaculation (complication occurred (1) versus no complication (0)), and re-operations (complication occurred (1) versus no complication (0)).

The input and output data were then arranged in a large table using Microsoft Excel.™ A highly abridged portion of the full input-output table is shown in Table 1.

**Table 1.** Example of the format in which the training data was organized

| **input** | | **desired output** | | |
|---|---|---|---|---|
| rhBMP2 (mg) | PDGF (ug) | fusion rate (%) | Oswesty score | New bone ($mm^2$) |
| 14 | 0 | 100 | 18.8 | . |
| 5 | 0 | 94.5 | 24.1 | . |
| 10 | 0 | 98.5 | 20.9 | . |
| 0 | 20 | . | . | 0.8 |
| 0 | 60 | . | . | 0.7 |
| 0 | 200 | . | . | 1.0 |

**Text S2: Deep neural networks**

The task at hand is to use machine learning to train some form of AI to extract the knowledge contained in an experimental input/desired-output dataset, and then use the AI to generalize on the basis of that knowledge to predict the outcomes of inputs on which it has not been trained. By far the most powerful AIs available for this kind of task are deep neural networks.[13,14] We evaluated several different network types in order to determine which type exhibited the best generalization capability for our dataset.

Deep neural networks are artificial neural networks that have potentially many processing layers. Like real neural networks, artificial neural networks are composed of many interconnected units, which can be thought of as highly simplified neurons. As in the real brain, the units can be organized into layers or circuits. Networks organized into layers are known as feedforward networks, while networks organized into circuits are known as recurrent networks.

The conceptually simplest connectivity pattern for feedforward networks is one in which every unit in a previous layer connects to every unit in the subsequent layer. The conceptually simplest connectivity pattern for recurrent networks is one in which every unit connects to every other unit, forming many circuits. In both cases this pattern is known as complete connectivity. All of the networks we evaluated had complete connectivity.

Each unit in an artificial neural network computes the sum of the activities of all the units that connect to it, after the activity of each sending unit is multiplied by the weight of its connection to the receiving unit. Thus, each unit computes its weighted input sum. The resulting activity of a linear unit will simply be its weighted input sum, unaltered by any further processing. The resulting activity of a nonlinear unit will be the weighted input sum after it has passed through a nonlinear activation function. The most common nonlinear activation function, and the one used here, is the sigmoidal activation function, which maps the real numbers into the range [0, 1]. Because any number of layers of linear units can be mathematically compressed into a single linear layer, all intervening layers in multi-layered neural networks should be nonlinear.

All neural networks have some units that are designed as input units, and others that are designated as output units. The activity levels of the input units are set from outside the network, while the activity levels of the output units are determined from the interactions among all the units in the network. The parameters of a neural network are the weights of the connections between the units. The weights are trained on pairs of input/desired-output patterns using machine learning, so that the network will produce the desired output pattern for each input pattern. The kind of machine learning in which a network is trained to produce a specific output for each input is known as supervised learning.

The simplest artificial neural network has only two layers: input and output; and is trained using the simplest form of supervised learning: the delta rule.[15] The input and output units can be linear. The next simplest artificial neural network has three layers: input, output, and hidden, where the hidden layer intervenes between the input and output layers. The hidden layer is so called because hidden unit activity is neither set (as for input units) nor trained (as for output units). The units in the hidden layer (as in all hidden layers) should be nonlinear.

Deep neural networks have more than one hidden layer and may have as many as ten or more hidden layers. Each additional hidden layer can add further processing power to an artificial neural network, and many layers are needed for certain applications. Artificial neural networks with any number of hidden layers can be trained using a generalization of the delta rule known as backpropagation.[15]

Recurrent neural networks have their units arranged in circuits, in which each unit can connect (and here does connect) with every other unit. Because the units can connect in many overlapping circuits, where each unit can both send to and receive from any other unit, recurrent networks process information in time. Recurrent networks are also deep networks, because every time step of processing in a recurrent network is equivalent to a layer in a feedforward network. Recurrent networks are trained using a generalization of backpropagation known as recurrent backpropagation.[16]

Most machine-learning algorithms have parameters that govern their operation. These parameters should be optimized for each network type to ensure the best training over the input/desired-output patterns in the dataset. The three supervised learning algorithms used here (delta-rule, backpropagation, and recurrent backpropagation) train network connection weights on the basis of the difference, or error, between the actual output and the desired output for any input. More specifically, they train the weights using weight-update terms that are derived from the gradient of the error with respect to the weights. The delta-rule, backpropagation, and recurrent backpropagation have two main parameters. The first is the learning rate, which is a positive constant (usually $< 1$) that is used to scale the weight-update terms. The second is the number of patterns presented to the network before their averaged weight-update terms are used to update the weights. This update-term averaging is known as stochastic gradient descent (SGD), and the number of terms in the SGD average can vary from 1 up to 100 or more.

A special type of feedforward neural network is the autoencoder, which is a neural network that is trained to reproduce the input as its desired output.[17] An autoencoder typically has one nonlinear hidden layer (but it could have several hidden layers) and is trained using backpropagation. The encoding of the input by an autoencoder hidden layer can have various uses, one of which is to serve as the input stage to a feedforward or recurrent neural network. Using an autoencoder encoding of the input, rather than using the raw input itself as the input to a neural network, can improve its performance. More specifically, using an autoencoder encoding as the input to a neural network can improve the ability of the neural network to generalize.

Neural networks are valuable as AIs because of their ability to *generalize* the knowledge they extract from a dataset to inputs that are not present in the dataset. Generalization enables a network to predict the outputs to inputs on which it has not been trained. Assessing the ability of a neural network to generalize is straightforward. To do so, the set of input/desired-output patterns is divided into a training set and a testing set (usually in a 75% - 25% split). Then the network is trained only on the input/desired-output patterns in the training set and is tested on the inputs in the testing set. Generalization error is quantified as the total root-mean-squared (RMS) error between the actual outputs and the desired outputs in the testing set. Networks with high generalization capability have low generalization error. Due to the randomness inherent in the machine-learning algorithms (delta-rule, backpropagation, and recurrent backpropagation), a proper generalization assessment involves retraining a network of a given type several times and averaging the RMS errors.

**Text S3: Finding the right neural network**

In principle there are no constraints on the number of neurons in a network, nor on the number of layers in a feedforward network, nor on the number of ways of constraining the connections in feedforward or recurrent networks, nor on any number of other neural network attributes. There is literally an infinite number of neural network types. Still, at least over a limited domain, care should be taken to ensure the suitability of the neural network for any specific application.

We evaluated 8 different neural network types, both with and without an autoencoder as a first stage, for a total of 16 different network configurations. The network types included a simple network having only an input layer and an output layer that was trained using the delta-rule. We called this network Delta. They also included feedforward networks with varying numbers of hidden layers that were trained using backpropagation. We called them BackOne, BackTwo, BackThree, BackFive, BackSeven, and BackTen, which had 1, 2, 3, 5, 7, or 10 hidden layers, respectively. They also included a recurrent network trained using recurrent backpropagation. We called this network Recur. In all networks the input and output units were linear while the hidden units were nonlinear, with their activations confined to the range [0, 1] using the sigmoidal nonlinear activation function.

Some initial evaluations using feedforward networks with 1 or 2 hidden layers showed good generalization capability when they had 100 hidden units per layer and were trained for 50,000 input/desired-output training pattern presentations. During training, patterns are chosen at random with replacement. With 50,000 training cycles, each of the 225 input/desired-output patterns in our dataset was presented to the network about 200 times. This number of training cycles was effective in reducing network error without overfitting (Figure S1).

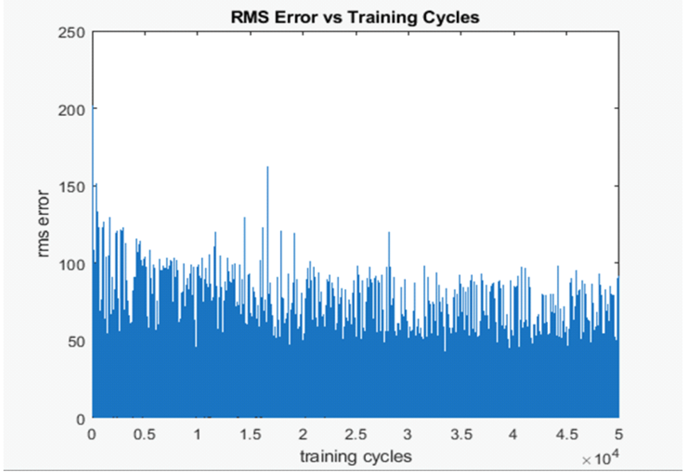


Figure S1. RMS error as a function of training cycles in a representative, feedforward neural network with 1 hidden layer of 100 nonlinear units. RMS error decreases as training proceeds and levels off after about 50,000 training cycles. Error remains relatively high because the dataset contains actual experimental results which differ between different labs, even for the same input values. Halting training after 50,000 training cycles provided adequate error reduction without overfitting the data, allowing good generalization capability.

As shown in Figure S1, RMS error decreases as training proceeds and levels off after about 50,000 training cycles. Since overtraining could compromise network generalization capability, there is no benefit in training beyond this point. It is clear from Figure S1 that RMS error stays relatively high even after 50,000 training cycles. The reason is that the dataset contains actual experimental results that vary between different labs. For example, the first five input/desired-output entries in the dataset all have exactly the same input values but have five different output values. Clearly, it would be impossible to reduce the error between the desired and actual outputs to zero in this case. Importantly, neural networks are excellent AIs to use in cases where there is ambiguity in the training data, because what they actually learn is to produce an output that is *probably* the best output, given the ambiguity in the dataset.[18]

We next optimized the training parameters for each network type. All hidden layers in feedforward networks had 100 hidden units. Also, there were 100 hidden units in the recurrent network, and it processed the input over 10 time steps. The parameters optimized were the learning rate ($a$) and the batch size, or the number of weight-update terms that were averaged before they were applied to the weights. The batch size is known as the stochastic gradient descent number (SGDnum) in supervised learning. Several rounds of adjustment in these parameters were needed to find the pair of $a$ and SGDnum that provided the greatest average error reduction over 10 retrainings of each network type. The results are shown in Table S1.

| **Optimized Parameters of All Network Types without Autoencoder** | | |
|---|---|---|
| **network type** | **learning rate ($a$)** | **Stochastic Gradient Descent (SGDnum)** |
| Delta | 0.01 | 1 |
| BackOne | 0.0025 | 1 |
| BackTwo | 0.01 | 7 |
| BackThree | 0.001 | 1 |
| BackFive | 0.001 | 2 |
| BackSeven | 0.00025 | 2 |
| BackTen | 0.0001 | 2 |
| Recur | 0.005 | 1 |

Table S1. Optimized learning parameters for all network types. Optimal learning rates ($a$) and the number of weight-update terms that were averaged before they were applied to update the weights (batch size or SGDnum) could vary considerably between network types.

Having optimized the parameters of our network types, we then assessed the ability of each to generalize. We divided the set of 225 input/desired-output patterns into 175 training patterns and 50 testing patterns. We then retrained each network type 10 times, choosing a different training set and testing set at random each time. The results are shown in Figure S2. They show that a feedforward neural network having one hidden layer generalizes the best.

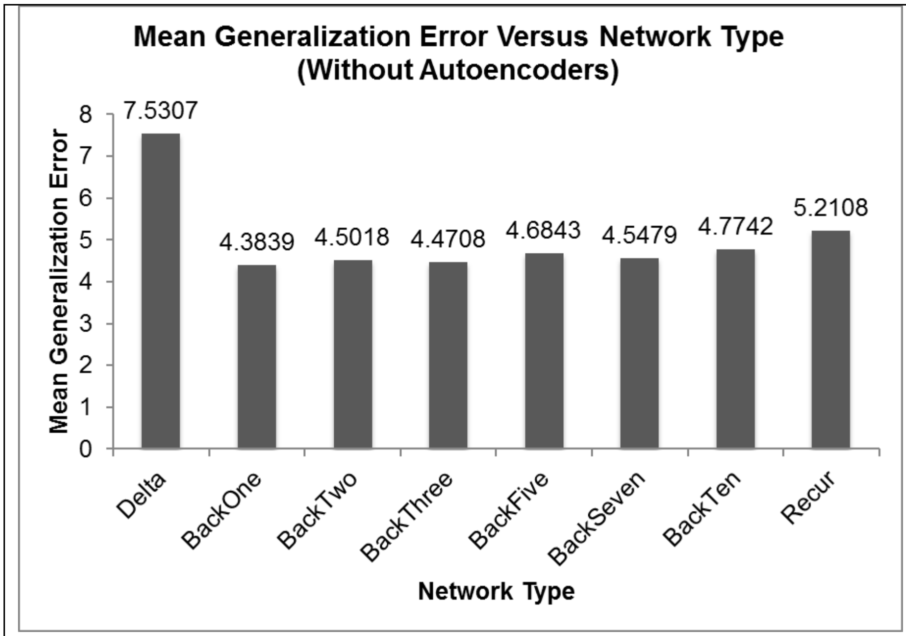


Figure S2. Assessing the ability of neural networks of different types to generalize. Networks of each type were each retrained and retested 10 times, with different input/desired-output patterns chosen each time for the training set and the testing set. The mean generalization error is the average over the 10 retrainings of the RMS error over the testing set. A feedforward neural network having one hidden layer generalizes the best.

We next trained an autoencoder using the input/desired-output values in the dataset. The autoencoder we used had one layer of nonlinear hidden units. We first optimized the training parameters (*a* and SGDnum) for autoencoders having different numbers of hidden units in the single hidden layer (5, 10, 20, 50, or 100). We then assessed the ability of an autoencoder having different numbers of hidden units to generalize over the dataset. Generalization was assessed as described above for the different neural network types. We found that an autoencoder having 50 hidden units showed the best generalization (data not shown).

In many applications, an autoencoder having fewer hidden units than inputs units (a so called undercomplete representation) will have the best generalization capability. Because our dataset has 17 inputs, it was expected that an autoencoder having 5 or 10 hidden units would generalize the best. However, the input in our dataset has three salient characteristics that make it less likely that an undercomplete representation will be optimal. The inputs in our dataset are: sparse, in that most input values are zero in most input patterns; noisy, in that the desired outputs for the same inputs can vary; and linear, so that different inputs can have different ranges of values (in our case some inputs range from 0 to 1 while others range from 0 to about 15). For these reasons it is not surprising that an overcomplete rather than an undercomplete representation had better generalization properties.[19] We found that an autoencoder having 50 hidden units generalized the best (data not shown).

We next optimized the training parameters for each of the network types with the autoencoded input representation rather than the raw input as the input to the network. The procedures for optimizing the training parameters for the network types with the autoencoder first stage were the same as those described above for the network types without the autoencoder. The results are shown in Table S2.

| **Optimized Parameters of All Network Types *With* Autoencoder** | | |
|---|---|---|
| **network type** | **learning rate (*α*)** | **Stochastic Gradient Descent (SGDnum)** |
| Delta | 0.05 | 1 |
| BackOne | 0.001 | 1 |
| BackTwo | 0.001 | 1 |
| BackThree | 0.00025 | 1 |
| BackFive | 0.0001 | 1 |
| BackSeven | 0.0005 | 1 |
| BackTen | 0.000025 | 1 |
| Recur | 0.01 | 1 |

Table S2. Optimized learning parameters for all network types with an autoencoder first stage. Optimal learning rates (*α*) varied widely between network types but all types seemed indifferent to the number of weight-update terms that were averaged before they were applied to update the weights (SGDnum). For simplicity, all SGDnum values were set to 1.

Finally, we assessed the ability of each network type with an autoencoder first stage to generalize. The procedure was the same as for the generalizability assessment described above without the autoencoder. The results are shown in Figure S3.

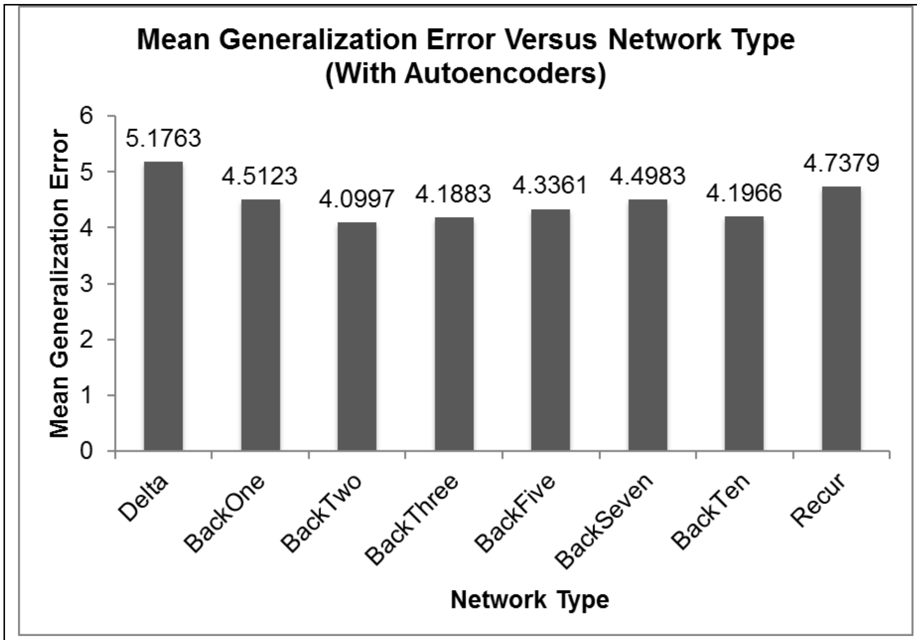


Figure S3. Assessing the ability of neural networks of different types, having an autoencoder as a first stage, to generalize. The generalization assessment was as described in Figure S2. A feedforward neural network with an autoencoder first stage and having two hidden layers generalizes the best. Note that the generalization error for this network is smaller than that for the feedforward network with one hidden layer that generalized the best without an autoencoder (see Figure S2).

With an autoencoder first stage, a feedforward network with two hidden layers showed the best generalization capability. The generalization error for this network is smaller than that for the feedforward network with one hidden layer that showed the best generalization capability for the network without an autoencoder first stage (Figure S2). The final result of these assessments is that the network type most suitable for our dataset is a feedforward neural network with two hidden layers, each with 100 hidden units, which also includes an overcomplete autoencoder of 50 hidden units as a first stage. A diagram of this network is shown in Figure 1 of the main text.

**Text S4: Possible implications of the character of the best network**

The fact that a feedforward neural network with three nonlinear layers (a nonlinear autoencoder layer and two nonlinear hidden layers) generalized best over the dataset could provide insight into the nature of the actual interactions among the biologic factors in the dataset. If all of the biologic factors acted independently, then the simplest network, having only linear input and output layers and trained using the delta rule, would have generalized as well as, if not better than, the more complex network types.

In the simplest network, each output is just a sum of the various inputs, after each input is multiplied by the weight of its connection to the output unit. The simplest network thus implements a linear combination of the inputs, and a linear combination would produce the best generalization in the case in which each input, which is 1 of the 17 different biologic factors, acts independently. But we found that generalization by the simplest network (Delta) was the worst among the network types we tested, whether with our without an autoencoder first stage.

The fact that generalization was the best in a neural network with three nonlinear layers interposed between the input and output layers indicates that the different orthobiologic factors do not act independently of one another, but instead interact in a complex way. It also suggests that the network was able to learn to represent at least part of the interaction between the different biologic factors, even though it was trained mainly on input/desired-output patterns in which the input included only one active factor. The possibility that the network did learn some of the interactions between the factors lends support to its predictions concerning the efficacies of combinations of the factors. This point is elaborated in the main text.

**Literature used to compile the training data:**


1. Aro, H.T., et al., *Recombinant human bone morphogenetic protein-2: a randomized trial in open tibial fractures treated with reamed nail fixation.* J Bone Joint Surg Am, 2011. **93**(9): p. 801-8.
2. Faundez, A., et al., *Bone morphogenetic protein use in spine surgery-complications and outcomes: a systematic review.* Int Orthop, 2016. **40**(6): p. 1309-19.
3. Gianakos, A., et al., *Bone Marrow Aspirate Concentrate in Animal Long Bone Healing: An Analysis of Basic Science Evidence.* J Orthop Trauma, 2016. **30**(1): p. 1-9.
4. Griffin, X.L., et al., *Electromagnetic field stimulation for treating delayed union or non-union of long bone fractures in adults.* Cochrane Database Syst Rev, 2011(4): p. CD008471.
5. Horstmann, P.F., et al., *(*) Composite Biomaterial as a Carrier for Bone-Active Substances for Metaphyseal Tibial Bone Defect Reconstruction in Rats.* Tissue Eng Part A, 2017. **23**(23-24): p. 1403-1412.
6. Jones, A.L., et al., *Recombinant human BMP-2 and allograft compared with autogenous bone graft for reconstruction of diaphyseal tibial fractures with cortical defects. A randomized, controlled trial.* J Bone Joint Surg Am, 2006. **88**(7): p. 1431-41.

7. Lee, D.H., et al., *Bone marrow aspirate concentrate and platelet-rich plasma enhanced bone healing in distraction osteogenesis of the tibia.* Clin Orthop Relat Res, 2014. **472**(12): p. 3789-97.
8. Marden, L.J., et al., *Platelet-derived growth factor inhibits bone regeneration induced by osteogenin, a bone morphogenetic protein, in rat craniotomy defects.* J Clin Invest, 1993. **92**(6): p. 2897-905.
9. Pocaterra, A., et al., *Effectiveness of platelet-rich plasma as an adjunctive material to bone graft: a systematic review and meta-analysis of randomized controlled clinical trials.* Int J Oral Maxillofac Surg, 2016. **45**(8): p. 1027-34.
10. Rutten, S., et al., *Enhancement of Bone-Healing by Low-Intensity Pulsed Ultrasound: A Systematic Review.* JBJS Rev, 2016. **4**(3).
11. Swiontkowski, M.F., et al., *Recombinant human bone morphogenetic protein-2 in open tibial fractures. A subgroup analysis of data combined from two prospective randomized studies.* J Bone Joint Surg Am, 2006. **88**(6): p. 1258-65.
12. Ye, F., et al., *Comparison of the use of rhBMP-7 versus iliac crest autograft in single-level lumbar fusion: a meta-analysis of randomized controlled trials.* J Bone Miner Metab, 2018. **36**(1): p. 119-127.

**Other References**

- LeCun Y, Bengio Y, Hinton G (2015) Deep Learning. *Nature* **521**: 436.
- Goodfellow I, Bengio Y, Courville A (2016) *Deep Learning*. MIT press.
- Rumelhart DE, McClelland JL, PDP Research Group (1986) *Parallel Distributed Processing. Explorations in the Microstructure of Cognition.* MIT Press.
- Pineda FJ (1989) Recurrent backpropagation and the dynamical approach to adaptive neural computation. *Neural Computation* **2**: 161.
- Hinton GE, Salakhutdinov RR (2006) Reducing the dimensionality of data with neural networks. Science **313**: 504.
- Richard MD, Lippmann RP (1991) Neural network classifiers estimate Bayesian a posteriori probabilities. *Neural Computation* **3**: 461.
- Vincent P, Larochelle H, Lajoie I, Bengio Y, Manzagol PA (2010) Stacked denoising autoencoders: Learning useful representations in a deep network with a local denoising criterion. *Journal of Machine Learning Research* **11**: 3371.
